# Effects of Opioid-free Anaesthesia on Postoperative Delirium after Gastrointestinal Surgery in Frail Elderly Patients: A Study Protocol for A Single-centre, Prospective, Randomized Controlled Trial

**DOI:** 10.64898/2026.08.12.26360246

**Authors:** Qiuyue Liu, Xiaoling Yang, Qiongyu Zhang, Min Zhang, Jianbo Wu, Yuehan Du, Yufei Li, Lina Chen, Xiaojun Gao, Yanyan Feng, Siyi Song, Xiaxuan Sun, Zekun Li, Lin Cheng, Yuyao Li, Mengjie Liu, Yongtao Sun

**Affiliations:** Department of Anesthesiology, Shandong Provincial Qianfoshan Hospital, Shandong Second Medical University, Shandong Institute of Anesthesia and Respiratory Critical Medicine, Shandong Provincial Clinical Research Center for Anesthesiology, Jinan, 250014, China; Department of Anesthesiology, The First Affiliated Hospital of Shandong First Medical University & Shandong Provincial Qianfoshan Hospital, Jinan, 250014, China; School of Anesthesiology, Shandong Second Medical University, Weifang, 261053, China; School of Clinical and Basic Medicine, Shandong First Medical University & Shandong Academy of Medical Sciences, Jinan, 250117, China; Department of Anesthesiology, Jinan Materrity and Child Care Affiliated to Shandong First Medical University, Jinan, 250000, China

**Keywords:** frailty, elderly patients, opioid-free anaesthesia, postoperative delirium, gastrointestinal surgery

## Abstract

**Introduction:** The incidence of postoperative delirium (POD) is high in frail elderly patients who have undergone gastrointestinal surgery, and POD significantly increases the risk of complications and medical burden. Opioid-free anaesthesia (OFA) involves a multimodal analgesic strategy, which may help to reduce the risk of POD. However, relevant studies focusing on frail elderly patients are still limited. This study aims to investigate the effect of OFA on the occurrence of POD in frail elderly patients after gastrointestinal surgery.

**Methods:** This single-centre, prospective, randomized controlled trial (RCT) will be conducted at the First Affiliated Hospital of Shandong First Medical University, China. A total of 174 frail elderly patients aged 65 years or older who plan to undergo elective gastrointestinal surgery (modified frailty index (mFI > 0.27) will be randomly assigned to either the OFA group (receiving dexmedetomidine, esmolol, and ketamine) or the opioid-based anaesthesia (OBA) group. The primary outcome is the incidence of POD within 7 days after surgery or at discharge. Secondary outcome measures include the perioperative stress response, inflammation, intraoperative haemodynamics, postoperative 30-day all-cause mortality, intraoperative haemodynamic changes, 15-item quality of recovery (QoR-15), and postoperative complications during hospitalization. This study focuses on frail elderly individuals (a high-risk population) and aims to investigate the potential benefits of the OFA strategy in reducing the incidence of POD by reducing exposure to opioids through multimodal analgesia. If positive results are obtained in this study, they may provide new evidence for optimizing the perioperative management of such patients. This study is a single-centre, prospective RCT, and the results can provide preliminary evidence for subsequent studies to be performed on a larger scale.

**Discussion:** This study focuses on frail elderly individuals (a high-risk population) and aims to investigate the potential benefits of the OFA strategy in reducing the incidence of POD by reducing exposure to opioids through multimodal analgesia. If positive results are obtained in this study, they may provide new evidence for optimizing the perioperative management of such patients. This study is a single-centre, prospective RCT, and the results can provide preliminary evidence for subsequent studies to be performed on a larger scale.

**Trial registration:** NCT07603596.

## Introduction

Postoperative delirium (POD) is a common and severe perioperative neuropsychiatric complication in elderly patients [1] and manifests primarily as acute and fluctuating impairment of consciousness, as well as cognitive dysfunction [2]. Previous studies have demonstrated that the occurrence of POD is closely related to a prolonged hospital stay, delayed functional recovery, long-term cognitive decline, and increased risk of death; moreover, POD has become an important clinical issue affecting the outcome of elderly surgical patients [3–6]. Gastrointestinal surgery is associated with severe trauma and a strong stress response, and the risk of POD in patients who undergo gastrointestinal surgery is greater than in those who undergo other types of surgery; thus, effective preventive strategies are urgently needed [7–9].

Frailty is a geriatric syndrome characterized by decreased physiological reserve and decreased stress tolerance, and the core festations of this condition include multisystem dysfunction and vulnerability to internal and external stimuli [3, 10]. Various tools, such as the Fried frailty phenotype (FFP) and the modified frailty index (mFI), have been widely used in frailty assessments, in which an mFI ≥0.27 has been proven to effectively predict adverse outcomes in elderly patients who have undergone surgery [11, 12].

Studies have demonstrated that the incidence of POD is significantly greater in frail patients than in nonfrail patients (54% vs. 20%), and the degree of frailty is positively correlated with the severity of POD [13–15]. The mechanism underlying the increased incidence of POD in frail patients may be related to decreased central nervous system reserves, excessive activation of inflammatory responses, and impaired blood–brain barrier function in these patients [3, 10]. In addition, pharmacokinetic abnormalities related to frailty further amplify the neurotoxic effects of perioperative interventions such as opioids.

Traditional perioperative analgesia relies on the use of opioids, which may increase the risk of POD by inhibiting the function of the prefrontal cortex, interfering with cholinergic neurotransmission, and disrupting the sleep–wake rhythm [16, 17]. Opioid-free anaesthesia (OFA) reduces opioid exposure through multimodal analgesic strategies (such as α2 receptor agonists, N-methyl-D-aspartate (NMDA) receptor antagonists, β blockers, and local anaesthesia techniques) and can theoretically reduce the incidence of POD. Although some studies have suggested that OFA can reduce opioid-related adverse reactions such as postoperative nausea and vomiting, its effect on POD is still controversial. Existing evidence has been obtained mostly from nonfrail populations or studies of mixed surgery types, and prospective studies targeting high-risk frail geriatric populations are particularly lacking. For example, a recent systematic review revealed that OFA can reduce the risk of POD during cardiac surgery; however, the sample exhibited high heterogeneity and was not corrected for frailty status. Therefore, it is necessary to further investigate the potential effect of OFA on POD in frail elderly patients undergoing gastrointestinal surgery through standardized randomized controlled trials (RCTs).

## Material and methods

### Study design

This single-centre, prospective randomized controlled trial is being conducted at the First Affiliated Hospital of Shandong First Medical University, China. Participant recruitment commenced on 1 May 2026. The study protocol was approved by the Ethics Committee of the First Affiliated Hospital of Shandong First Medical University (Approval No. YXLL-KY-2025 (079)). Written informed consent will be obtained from all participants before enrolment. All study personnel have received training in the ethical conduct of human participant research. The protocol complies with the Standard Protocol Items: Recommendations for Interventional Trials (SPIRIT) statement [18]. A total of 174 participants will be randomly assigned to the opioid-based anaesthesia (OBA) group or the OFA group at a ratio of 1:1. The timetables for subject enrolment, study interventions, and outcome evaluation after the SPIRIT statement are shown in **Fig. 1**. The flowchart of this study is shown in **Fig. 2**.

**Fig. 1.**
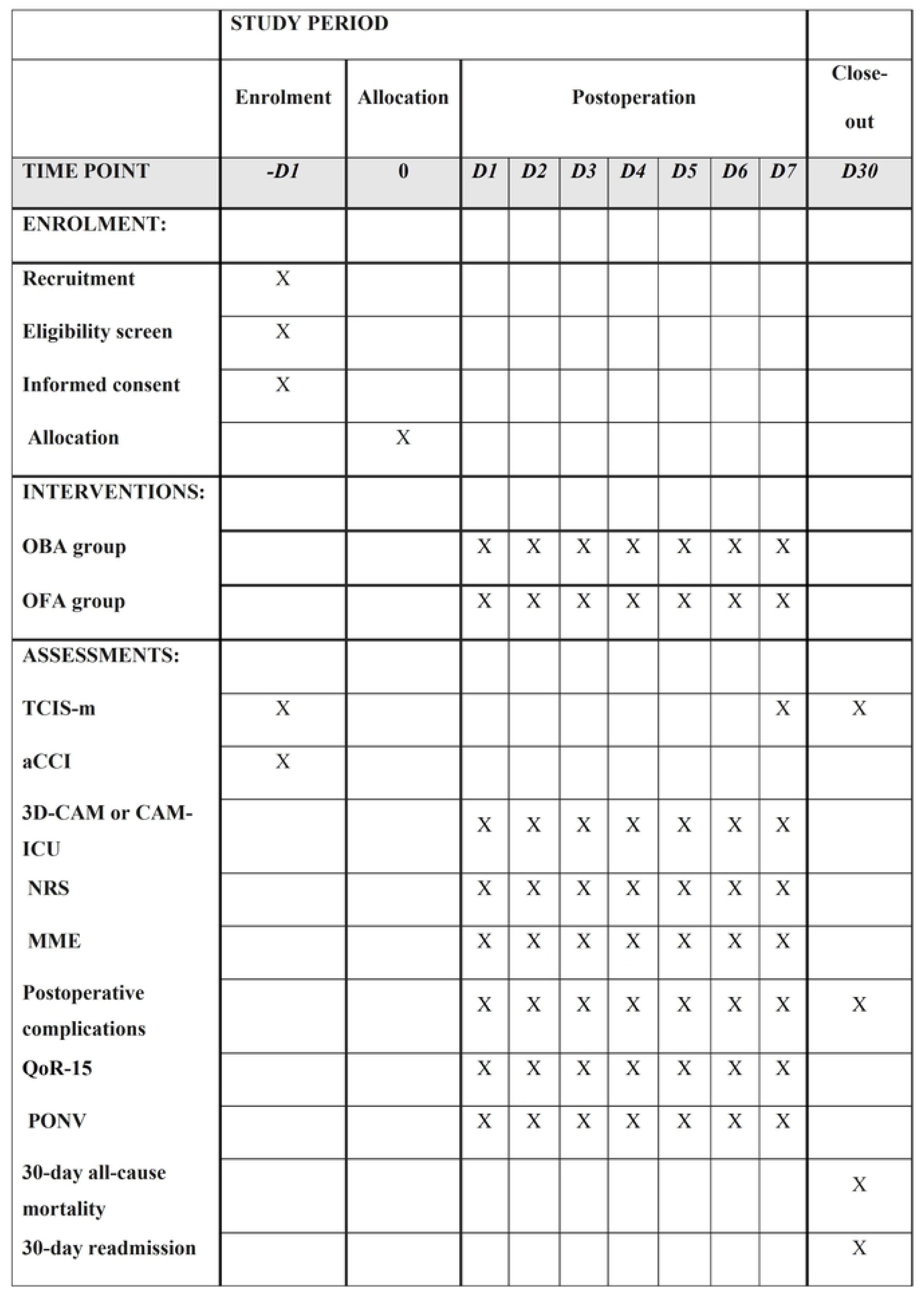
Timeline of the study. OBA group, opioid-based anaesthesia group; OFA group, Opioid-free anaesthesia group; TICS-m, Telephone Interview for Cognitive Status-modified; aCCI, Age-adjusted Charlson Comorbidity Index; 3D-CAM, 3-Minute Diagnostic Interview for CAM; NRS, Numerical Rating Scale; MME, Morphine Milligram Equivalent; QoR-15, 15-item quality of recovery; PONV, Postoperative nausea and vomiting.

**Fig. 2.**
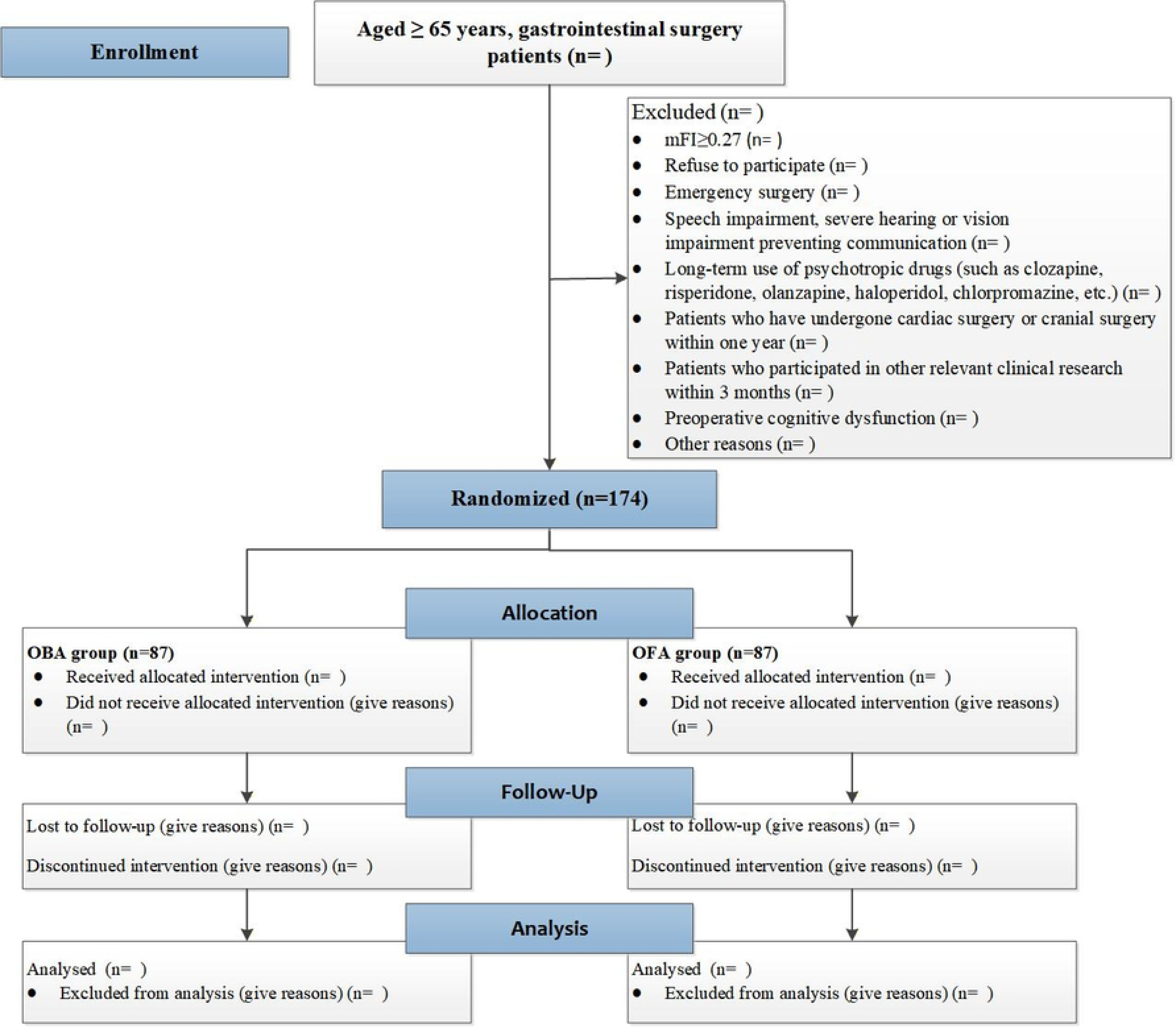
Flow diagram of the study.

### Trial objectives

The main objective of this study is to investigate the effect of OFA on POD in frail elderly patients after gastrointestinal surgery.

### Study subjects

Frail elderly patients (aged ≥65 years) who plan to undergo elective gastrointestinal surgery under general anaesthesia will be selected.

### Inclusion criteria

1. Patients who plan to undergo elective gastrointestinal surgery under general anaesthesia;
2. Age ≥65 years;
3. American Society of Anaesthesiologists (ASA) grade I–III;
4. Body mass index (BMI) of 18.0–30.0 kg/m^2^;
5. Frailty: mFI ≥ 0.27;
6. Signed informed consent.

### Exclusion criteria

1. Patients undergoing emergency surgery;
2. Patients with language disorders or severe hearing or vision impairment that prevent communication;
3. Patients with a history of neurological and psychiatric disorders, including Alzheimer’s disease (AD), other types of dementia, stroke, and psychosis;
4. Patients with long-term use of psychotropic medications (such as clozapine, risperidone, olanzapine, haloperidol, and chlorpromazine);
5. Patients who have undergone cardiac surgery or craniocerebral surgery within the past year;
6. Patients who have participated in other relevant clinical trials within the past 3 months;
7. Patients with preoperative cognitive impairment (Telephone Interview for Cognitive Status-modified (TICS-m) score ≤ 27), as determined via the TICS-M test;
8. Each patient can be only included only once, regardless of whether the reason for the second surgery is related to the primary cause.

### Shedding criteria

1. Tests are not performed according to the research requirements;
2. Serious adverse events (SAEs) occur during the trial;
3. Patients request withdrawal during the study;
4. Study participants are lost to follow-up;
5. Participants lack study records or have incomplete records for other reasons.

The detailed reasons for the withdrawal of participants are documented in the case report form (CRF). If a participant voluntarily withdraws from the study, the investigator should ask and record the reason for the withdrawal. If a study participant withdraws from the study because of an adverse event (AE), the investigator should follow up on the AE until it is resolved or stabilized.

### Study implementation

All members of the research team will receive systematic training before the study and be proficient in administering and interpreting the TICS-m [19], the 3-Minute Diagnostic Interview for CAM (3D-CAM) [20], the Confusion Assessment Method for the Intensive Care Unit (CAM-ICU) [21], morphine milligram equivalent (MME) calculation [22], vital signs data collection, the Numerical Rating Scale (NRS) for Pain [23], and the 15-item Quality of Recovery Index (QoR-15) [24]. A detailed explanation of the perioperative evaluation indicators is provided in Supplement 1.

### Randomization and blinding

This study will use block randomization. Stata software will be used to generate blocks. The block sizes will be 2, 4, and 6, which randomly will change. Included patients will be randomly assigned to the OBA group or the OFA group at a 1:1 ratio. This study is a single-blind trial; specifically, the study participants will be blinded, but the anaesthesiologists will not be blinded to ensure the smooth implementation of intraoperative anaesthesia management. The grouping information will be sealed in a sealed opaque envelope and sequentially numbered. When a patient enters the operating room, the anaesthesiologist will open the corresponding numbered envelope and determine the patient group according to the grouping information written on the card inside of the envelope. Patients, surgeons, nurses, postoperative visitors, and statisticians will remain blinded to the group information. Postoperative follow-up and evaluation of study outcomes will be conducted by independent investigators to minimize bias associated with data collection.

### Study interventions

#### General anaesthesia and analgesia protocol

All patients will undergo general anaesthesia with tracheal intubation or a laryngeal mask airway. No preoperative anaesthesia medication will be administered, and the patient will directly enter into the operating room. Routine monitoring items will include electrocardiogram (ECG), pulse oximetry (SpO_2_), blood pressure, partial pressure of end-tidal carbon dioxide (P_ET_CO_2_), and anaesthesia depth (monitored by using the bispectral index (BIS)). Invasive blood pressure or central venous pressure monitoring will be performed as needed. Prior to anaesthesia induction, the patient will be preoxygenated via inhalation of 100% oxygen through a mask for 5 min. Anaesthesia induction, intraoperative maintenance, and postoperative analgesia will be performed for the two groups of patients according to the following schemes.

In the OBA group, anaesthesia will be induced with 1.5–2.5 mg/kg propofol, 0.6–1.0 mg/kg rocuronium, and 0.3–0.5 μg/kg sufentanil. Endotracheal intubation or laryngeal mask airway insertion will be performed when the BIS value decreases to 40–60. During surgery, 2–6 mg/kg/h propofol, 0.2–1.0 μg/kg/min remifentanil, and 5–6 μg/kg/min rocuronium will be continuously pumped to maintain anaesthesia. Both groups will receive inhalation of 1% sevoflurane during anaesthesia maintenance. At the end of the procedure (after the last suture), all maintenance drugs will be discontinued. Afterwards, 8 mg of ondansetron and 0.15 μg/kg sufentanil will be intravenously injected. Postoperative analgesia will be performed by using an intravenous patient-controlled analgesia pump; the formulation will involve 2 μg/kg sufentanil and 16 mg of ondansetron, which will be diluted to 100 mL with 0.9% normal saline. Moreover, the background infusion rate will be set at 2 mL/h, and the lockout interval will be 15 min.

In the OFA group, anaesthesia will be induced with 1.5–2.5 mg/kg propofol, 0.3–0.5 mg/kg esketamine, and 0.6–1.0 mg/kg rocuronium. Endotracheal intubation or laryngeal mask placement will be performed when the BIS value is observed at 40–60. Intraoperative continuous intravenous infusion of 2–6 mg/kg/h propofol, 0.5–1.0 μg/kg/h dexmedetomidine, 0.2–0.5 mg/kg/h esketamine, 5–6 μg/kg/min rocuronium bromide, 20–100 μg/kg/min esmolol, and inhaled 1% sevoflurane will be utilized to maintain anaesthesia. At the end of the procedure (after the last suture), all maintenance drugs will be discontinued. Afterwards, 8 mg of ondansetron and 0.2 mg/kg esketamine-assisted sufentanil will be intravenously administered. Postoperative analgesia will be performed by using an intravenous patient-controlled analgesia pump consisting of 1.5 mg/kg esketamine and 16 mg of ondansetron. The volume will be adjusted to 100 mL in 0.9% normal saline; moreover, the continuous infusion rate will be set at 2 mL/h, and the lockout interval will be 15 min.

The following respiratory parameters will be established for both groups: tidal volume of 6–8 mL/kg, ventilation frequency of 10–14 breaths per minute, inspiration to expiration ratio of 1:1.5–2, airway pressure <30 mmHg, and positive end-expiratory pressure (PEEP) of 5–0 cmH₂O, with the goal to maintain the P_ET_CO_2_ at 35–45 mmHg. After anaesthesia is induced, bilateral ultrasound-guided transversus abdominis plane block will be performed by using 40 mL of 0.375% ropivacaine. If haemodynamic instability occurs during surgery (e.g., a systolic blood pressure <90 mmHg or >160 mmHg and a heart rate <50 beats/min or >100 beats/min), this event will be recorded as an AE. Events will be classified as SAEs only if organ dysfunction, prolonged hospital stay, or life support intervention occurs. After the operation, the patients will be transported to the postanaesthesia care unit (PACU) and intravenously administered sugammadex sodium at a dose of 2–4 mg/kg to reverse the neuromuscular blockade.

### Outcomes Primary outcome

Incidence of POD within 7 days after surgery or at discharge.

### Secondary outcomes

a. Serum inflammatory factor concentrations (including serum tumour necrosis factor-α (TNF-α) and interleukin-6 (IL-6)) and perioperative stress responses (including cortisol, epinephrine, and norepinephrine serum concentrations). Serum samples will be collected at the following time points: 10 min after anaesthesia induction; 10 min and 1 h after pneumoperitoneum; and 24 h and 48 h after surgery.
b. All-cause mortality within 30 days after surgery.
c. Intraoperative haemodynamic changes recorded at the following time points: before tracheal intubation; immediately after tracheal intubation; at the beginning of the operation; at 30 min (± 5 min), 60 min (± 5 min), 120 min (± 5 min), and 5 min after pneumoperitoneum; at the end of the operation, and at extubation.
d. The QoR-15, which will be assessed 24 h, 48 h, and 72 h after surgery.
e. Postoperative complications during hospitalization (such as pulmonary infections, urinary tract infections, cardiovascular and cerebrovascular accidents, abnormal liver function, postoperative bleeding, incisional infections, deep vein thrombosis in the lower extremities, electrolyte imbalances, hypoalbuminaemia, incisional infections, wound site infections, and postoperative bleeding).
f. The NRS pain score, which will be evaluated at 72 h after surgery.
g. Incidence of nausea and vomiting within 72 h after surgery.
h. The MME of analgesics at 72 h after surgery.
i. Duration of anaesthesia (from the induction of anaesthesia to the withdrawal of anaesthetic medications).
j. Operation time (from skin incision to the last suture).
k. Length of PACU stay.
l. Length of hospital stay.

### Data management and safety monitoring

All evaluations will be performed by members of the study team who are unaware of treatment grouping. All of the investigators will receive standardized training on neurocognitive and delirium assessments before the initiation of the study. Missing intraoperative data will be obtained from electronic medical records. Postoperative data will be obtained through interviews and electronic medical records within 7 days after surgery. The Ethics Committee of the First Affiliated Hospital of Shandong First Medical University will complete the monitoring of research and data quality. We do not expect this study to expose the participants to any serious risk. All of the AEs (regardless of whether they are related to the trial drugs) will be monitored and recorded. The anaesthesiologist will decide whether to terminate the trial based on the severity of the AEs. All of the AEs will be promptly treated, and the reasons for the temporary termination of the trial will be recorded.

### Sample size calculation

The primary outcome of this study is the incidence of POD within 7 days after surgery or before discharge. Based on the literature review and the previous research results of our center [25–27], the incidence of POD in elderly frail patients undergoing gastrointestinal surgery is estimated to be 54.8% in the opioid-based anesthesia (OBA) group and 32.6% in the opioid-free anesthesia (OFA) group. The two-tailed test level is set at α = 0.05, the power of the test is 1-β = 0.80, and the sample size ratio of the two groups is 1:1. As determined using PASS 2021 software, approximately 78 frail patients in each group (a total of 156 cases) are needed. Accounting for an anticipated 10% loss to follow-up or incomplete data, the final plan includes 87 patients per group, resulting in a total sample size of 174 elderly frail patients undergoing gastrointestinal surgery.

### Statistical analysis

All of the statistical analyses will be performed by using SPSS software and will follow the intention-to-treat (ITT) principle; specifically, all of the randomized, grouped patients will be included in the final analysis. Continuous variables will be expressed as the mean ± standard deviation or median (interquartile range) according to their distribution, and categorical variables will be expressed as frequencies and percentages. The normality test will be performed by using the Shapiro-Wilk test or the Kolmogorov-Smirnov test.

Analysis of the primary outcome: The primary outcome is the incidence of POD within 7 days after surgery or before discharge. The incidence of POD between the two groups will be compared by using the chi-square test or Fisher’s exact test, and the relative risk (RR) and its 95% confidence interval (CI) will be calculated. If there is an imbalance observed between the groups regarding the baseline data, a multivariate logistic regression model will be used for correction analysis; however, this analysis only involves an exploratory study.

Analysis of secondary outcomes: Appropriate statistical methods will be selected according to the data type. Continuous variables (such as QoR-15 scores, NRS pain scores, length of PACU stay, length of hospital stay, and opioid consumption) will be compared between the groups by using independent samples t tests (for normally distributed variables) or Mann-Whitney U tests (for nonnormally distributed variables). Categorical variables (e.g., postoperative nausea and vomiting, incidence of complications, intensive care unit (ICU) admission rate, 30-day readmission rate, and mortality) will be analysed by using the chi-square test or Fisher’s exact test. Repeated measures data (such as inflammatory cytokine levels, haemodynamic indicators, and pain scores detected at different time points) will be analysed by using repeated measures analysis of variance (ANOVA) or generalized estimating equations (GEEs) to evaluate the differences between the groups and the time interaction effect.

Processing of missing data: The multiple imputation method will be used to process the missing data of the major outcome indicators; additionally, the pattern and proportion of the missing data will be reported.

Significance level: All of the statistical tests will involve two-tailed tests, and P < 0.05 will be considered to indicate statistical significance.

### Hazard

AEs will be recorded from baseline (at the beginning of the study) to two weeks after the last follow-up (FU12), regardless of whether there is a causal relationship detected between the interventions. Adverse behaviours and SAEs will be recorded in detail during the intervention and within two weeks after each assessment visit. The severity of AEs will be classified into grades 1 to 5: mild (grade 1), moderate (grade 2), severe (grade 3), life-threatening (grade 4), and fatal (grade 5). If the SAE is rated as grade 2 or above, it will be reviewed and verified by the medical staff.

All SAEs (> grade 2) reported during follow-up and AEs (grade > 1) reported during the intervention period will be reviewed and verified by the independent Data Safety Monitoring Board (DSMB) to verify their causality and severity classification. The DSMB will discuss and decide on changes to or termination of the interventions based on the SAE report. All of the participants will be covered by insurance and travel accident insurance.

### Proposal revision

During the implementation of the study, the study protocol may require revision because of practical operations, ethical requirements, or scientific progress. All of the protocol revisions will follow certain procedures. Specifically, any modifications of the study design, target population, intervention measures, outcome measures, or statistical methods must be proposed by the person in charge or the principal investigator of the study; moreover, the reason for the revision and the expected impact should be explained. All of the revisions must be submitted to the Ethics Committee of the First Affiliated Hospital of Shandong First Medical University for review and approval before implementation. The key contents of the protocol may not be altered without the approval of the ethics committee. If the revision involves important changes in the primary outcome, inclusion and exclusion criteria, or interventions, the registration information will be promptly updated at ClinicalTrials.gov (NCT07603596) to ensure the transparency of the study. If the revisions may affect the subjects’ rights, safety, or willingness to participate, written informed consent from the subjects or their legal representatives should be obtained once more. All protocol revisions, dates, reasons, and approval documents will be fully archived and explained in the final study report. This study is committed to strictly following ethical norms and scientific principles to ensure compliance and transparency in the protocol revision process.

## Discussion

This study mainly investigates the effect of OFA on the incidence of POD in frail elderly patients after gastrointestinal surgery. Frail elderly patients exhibit a high risk for POD [16, 28]; however, few OFA studies have targeted this population [29, 30].

The main characteristics of this study are as follows. First, this study focuses on frail elderly individuals (a high-risk population); moreover, the use of mFI ≥ 0.27 as the screening criterion represents a more targeted measure than age stratification alone. Frail patients exhibit greater sensitivity to opioid medications and may theoretically derive greater benefits from the OFA strategy [31, 32]. Second, the OFA regimen (involving the combination of dexmedetomidine, esketamine, and esmolol) can avoid the use of opioids while achieving analgesic and sedative effects. These three drugs may synergistically reduce the risk of POD through various pathways, such as anti-inflammation and the stabilization of blood flow [33, 34].

We hypothesize that the incidence of POD in the OFA group will be significantly lower than that in the traditional OBA group. The mechanism of this effect is based on the fact that opioids promote POD through various pathways, such as by activating neuroinflammation, interfering with cholinergic transmission, and suppressing respiration [35]. In contrast, the OFA strategy may mitigate central neuroinflammation and oxidative stress damage by avoiding opioid exposure and being combined with organ-protective drugs [33]. Secondary outcome measures (such as inflammatory cytokine levels, stress hormone levels, pain scores, and recovery quality) can help to elucidate the underlying mechanisms [28, 36].

In addition, secondary outcome measures include complication rate, 30-day mortality, length of hospital stay, haemodynamic stability, and opioid consumption [29, 37]. The comprehensive evaluation of these indicators can help to determine whether the OFA strategy can result in more extensive clinical benefits, such as promoting postoperative rehabilitation and reducing opioid-related adverse reactions, in addition to reducing POD [33, 38]. For frail elderly patients, any interventions that can reduce complications and shorten hospital stays demonstrate important health and economic values [39, 40].

This study has several limitations. First, it utilizes a single-centre and small-sample design, and the conclusions should be extrapolated with caution. Second, anaesthesiologists cannot be blinded; however, the outcome evaluators and statisticians will be blinded to the results to minimize bias. The combined application of multiple medications requires further investigation of their interactions and optimal compatibility.

## Conclusion

In summary, this study is a prospective RCT designed to provide preliminary evidence for the application of the OFA strategy in gastrointestinal surgery for frail elderly patients. The results of this study may provide the foundation for larger-scale deterministic studies in the future and may demonstrate promise for providing optimized strategies for perioperative management in this high-risk population.

## Data Availability

No datasets were generated or analysed during the current study. All relevant data from this study will be made available upon study completion.

## Acknowledgments

We would like to thank the staff at the Center for Big Data Research in Health and Medicine, The First Affiliated Hospital of Shandong First Medical University and Shandong Provincial Qianfoshan Hospital for their valuable contributions.

## Consent for publication

Not applicable.

## Availability of data and materials

Not applicable.

## Author Contributions

Qiuyue Liu: Methodology, Software, Validation, Formal analysis, Writing-original draft.

Xiaoling Yang: Conceptualization, Supervision, Software, Investigation, Project administration, Writing-original draft.

Qiongyu Zhang: Methodology, Software, Validation, Formal analysis, Writing-original draft.

Jianbo Wu: Conceptualization, Supervision, investigation, Resources, Data curation, Writing-review & editing.

Yuehan Du: Investigation, Data curation, Writing-original draft.

Min Zhang: Validation, Formal analysis, Project administration, Writing-original draft. Yufei Li: Software, Investigation, Project administration, Writing-original draft.

Lina Chen: Conceptualization, Supervision, Software, Investigation, Project administration, Writing-original draft.

Xiaojun Gao: Investigation, Resources, Data curation, Writing-original draft. Yanyan Feng: Software, Investigation, Project administration, Writing-original draft. Siyi Song: Software, Investigation, Visualization, Writing-original draft.

Xiaxuan Sun: Validation, Formal analysis, Writing-original draft. Zekun Li: Investigation, Project administration, Writing-original draft.

Lin Cheng: Investigation, Resources, Data curation, Writing-original draft. Yuyao Li: Software, Investigation, Visualization, Writing-original draft.

Mengjie Liu: Software, Investigation, Visualization, Writing-review & editing. Yongtao Sun: Conceptualization, Methodology, Data Curation, Project administration, Funding acquisition, Writing-review & editing.

